# Acute intermittent hypoxia in people living with chronic stroke – a preliminary study to examine safety and efficacy as a neurorehabilitation intervention

**DOI:** 10.1101/2023.12.01.23299309

**Authors:** Gregory EP Pearcey, Alexander J Barry, Milap S Sandhu, Timothy Carroll, Elliot J Roth, W Zev Rymer

## Abstract

**Background and Purpose:** Acute intermittent hypoxia (AIH) is a novel therapeutic intervention that has the potential to facilitate recovery of function, but its safety and efficacy have not been tested in people with stroke. The purpose here was to examine whether AIH is safe and effective in people with stroke.

**Methods:** Participants (n=10) with a unilateral, ischemic, hemispheric stroke were assessed before and following 4 sessions of AIH. Clinical tests and upper limb strength were assessed before, ∼15-30 minutes, and ∼60 minutes after the intervention.

**Results:** AIH was well-tolerated and there were no adverse events observed. Although no changes in strength were detected for the less-affected limb, grip strength and elbow flexion force of the more-affected limb was increased after AIH.

**Conclusions:** AIH appears to be potentially safe and effective for improving strength in the more-affected limb in people with stroke. Future work should explore the use of AIH to enhance task-specific training-induced plasticity.

## INTRODUCTION

Stroke is a leading cause of long-term disability worldwide. Despite some spontaneous recovery following a stroke, many people with stroke show substantial residual functional impairments that significantly affect their independence and quality of life. Novel interventions to alleviate impairment in people with stroke are urgently needed. Several therapies promote strength and endurance, but few capitalize on the unique potential of the central nervous system to reorganize following injury.

Acute intermittent hypoxia (AIH), a protocol that involves brief respiratory exposure to reduced oxygen concentrations, has been found to augment neuroplasticity and promote functional recovery after spinal cord injury (SCI)^1^. Although the pathology of stroke differs from SCI, stroke is often accompanied by damage to white matter projections. Therefore, there is great potential for similar therapeutic benefits arising from transient reductions in oxygen on damaged cerebral projections. Published results in people with SCI have shown enhanced strength and coordination rather quickly after AIH, which opens a temporary window for enhanced neuroplasticity after a single intervention^2^.

The purpose of this study was to investigate the safety and efficacy of AIH in people with stroke. The objectives were two-fold, to examine whether brief reductions in inhaled oxygen 1) are safely tolerated, and 2) improve upper limb strength. We hypothesized that AIH would be safely tolerated and increase strength on the more affected side.

## METHODS

### Participants

Sixteen people were recruited and consented to complete the study, but only 10/16 completed all procedures. Data are thus reported on 10 individuals (58.2±5.8 years, 2 females) living with chronic stroke (10.4±7.1 years since injury). The remaining 6 participants were either withdrawn due to resting heart rates <50 bpm, dropped out due to positive tests for Covid-19, or did not qualify following the screening session. The experimental protocol was registered (NCT04019522) and participants provided informed and written consent to procedures approved by the local ethics committee at Northwestern University (IRB protocol STU00208610).

### Experimental sessions

Participants attended a free-standing academic rehabilitation hospital for six visits. The first visit was a baseline assessment, which included a structural magnetic resonance imaging (MRI) scan; a medical history, demographics, and medications; a pregnancy test for women of childbearing potential; performance of the outcome assessments; medical monitoring; and a blood draw. The next four sessions had a consistent format, but the level of inspired oxygen during AIH was modified from 21% (i.e., room air) in the first session, reduced by 4% each session to reach 9% in the final experimental session. In each experimental session, participants performed outcome assessments before and after 15 sequences of brief oxygen reductions while vital functions were monitored. After AIH, blood was drawn, and a neurological assessment was performed by a trained physician. Each visit was separated by at least 48 hours. The final visit was a follow-up assessment, which included a structural MRI scan; performance of the outcome assessments; medical monitoring; and a blood draw.

### AIH intervention

Each session utilized a customized Hyp-123 device (Hypoxico, Gardiner, NY) to switch between hypoxic air and normoxic air. Participants breathed lowered O_2_ concentrations for 30-60 seconds, followed immediately by breathing room air for 60-90 seconds, and this was repeated for 15 cycles. The total duration of each intervention was approximately 30 minutes and O2 concentrations were monitored with a Maxtec Handi+ oxygen analyzer (Maxtec, Salt Lake City, UT).

Since this was the first study using the AIH protocol in people with stroke, a titration approach was used for the O2 concentrations across sessions and was decreased along the above-mentioned schedule. This allowed the capacity for monitoring participants for adverse events while slowly increasing the hypoxia levels to identify any potential issues. A side effect questionnaire was given before, during, and after AIH where participants were instructed to inform study staff if they experienced any notable changes.

### Physiological monitoring

Participants oxygen saturation levels (SpO2) and heart rate were monitored with a Nonin 7500 SpO2 monitor (Nonin Medical Inc., Plymouth, MN), and a 5-lead electrocardiogram was read in real time by a registered nurse. Blood pressure was measured before, during, and after the interventions.

### Outcome Assessments

#### Clinical assessments

At their first visit, all participants underwent a battery of clinical assessments, prior to AIH administration. A medical history was taken by the physician to confirm that the participant met inclusion and exclusion criteria and that there were no other co-morbidities. Participants underwent a structural brain MRI before the intervention to confirm size and location of the stroke lesion, and after the intervention to determine whether the stroke lesion size changed. Participants then received a 12-lead EKG assessment at rest, which the study physician used to identify whether cardiac abnormalities were present prior to intervention. A baseline blood draw was assessed for Troponin levels, and a second blood draw was assessed for Troponin levels after each intervention session, before each participant was discharged.

Prior to and following each intervention, a trained clinician performed: neurological assessments to determine whether there were changes in cognition (NIH Stroke Scale, a measure of stroke severity); cranial nerve assessment; a standard muscle strength test; the Brunnstrom scale to assess level of motor recovery; sensory changes; reflexes; coordination; assessment of heart and lung status; Fugl-Meyer test, a quantitative measure of motor impairment; Chedoke McMaster Stroke Assessment, a measure of impairment of the hand; Modified Ashworth Scale for Spasticity, and Delis Kaplan Executive Function System Color-Word Interference Test, an assessment of verbal processing and executive functioning.

#### Strength testing

Elbow flexion strength was assessed using a custom elbow dynamometer that utilizes a TAS510 tension/compression load cell (HTC Sensors, Xi’An, China) to quantify elbow flexion force. Grip strength was measured using a Jamar analog hand dynamometer (Jamar Technologies, Hatfield, PA). For both elbow flexion and grip strength, the peak force obtained during a three second maximal voluntary contraction was recorded. w60 seconds rest was provided before the next trial. A total of three trials of each test was performed in each limb. The right limb was tested first, followed by the left limb and the mean of three trials was used for further analysis.

## RESULTS

### Safety

Our primary objective was to determine if AIH is safely tolerated by people with stroke. Across participants and sessions, there were no adverse consequences observed in clinical assessments, neurological assessments, or troponin levels after AIH. Participants tolerated breathing 9% O2, which caused SpO2 to drop to a nadir of 86.9±0.9% (P<0.001; fig.1A) and heart rate to increase by 7.3±4.8 beats per minute (P<0.001; fig.1B). Significant differences in apparent diffusion coefficient (ADC) values were observed in the MRI scans but secondary analysis found that voxels with drops in ADC were disjoint and located on the periphery of the brain. Thus, they were deemed to be a result of imperfection in the image registration (i.e., noise), further excluding negative consequences in brain morphology because of the AIH intervention.

**Figure 1:**
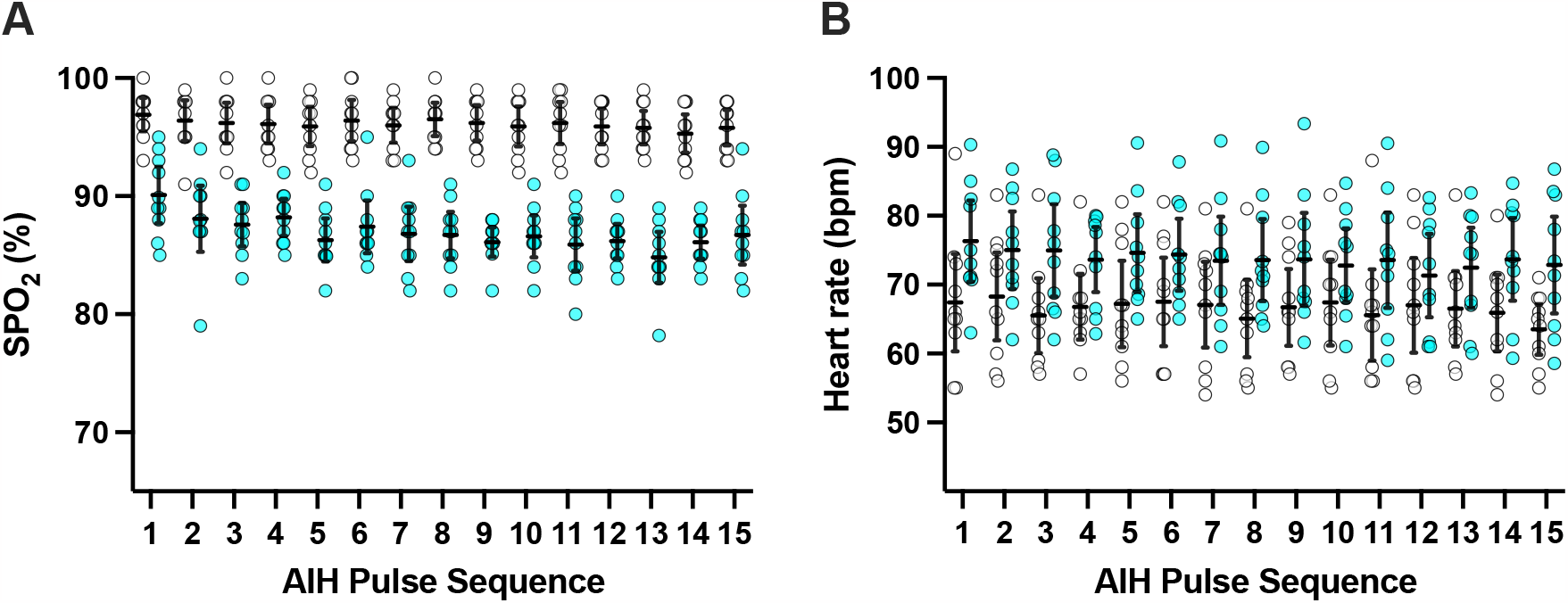
Physiological monitoring during acute intermittent hypoxia (AIH). A) the nadir oxygen saturation reached during each sequence of AIH. B) the maximum heart rate reached during each sequence of AIH. In all plots, the 21% O2 condition is in white, while the 9% O2 condition is in cyan. Error bars are 95% confidence limits. All timepoints were significantly different between conditions in A and B.

### Strength

The secondary objective was to determine if AIH improves strength in the upper limb of stroke survivors. To account for within-participant variability, we computed the percent change from baseline during the breathing of 21% and 9% O2 (AIH). The change in elbow flexion strength in the more affected limb during the 9% O2 condition (4.92±3.34% *increase* from baseline) was significantly greater than the change during the 21% O2 condition (7.6±3.34% *decrease* from baseline; p=0.003; fig.2A). Similarly, in the more affected limb, the change in grip strength during the 9% O2 condition (12.9±6.38% *increase* from baseline) was significantly greater than the change during the 21% O2 condition (12±3.81% *decrease* from baseline; p=0.012, fig.2C). The change in grip strength and elbow flexion strength from baseline was not different between conditions in the less affected limb (fig.2B and 2D).

**Figure 2:**
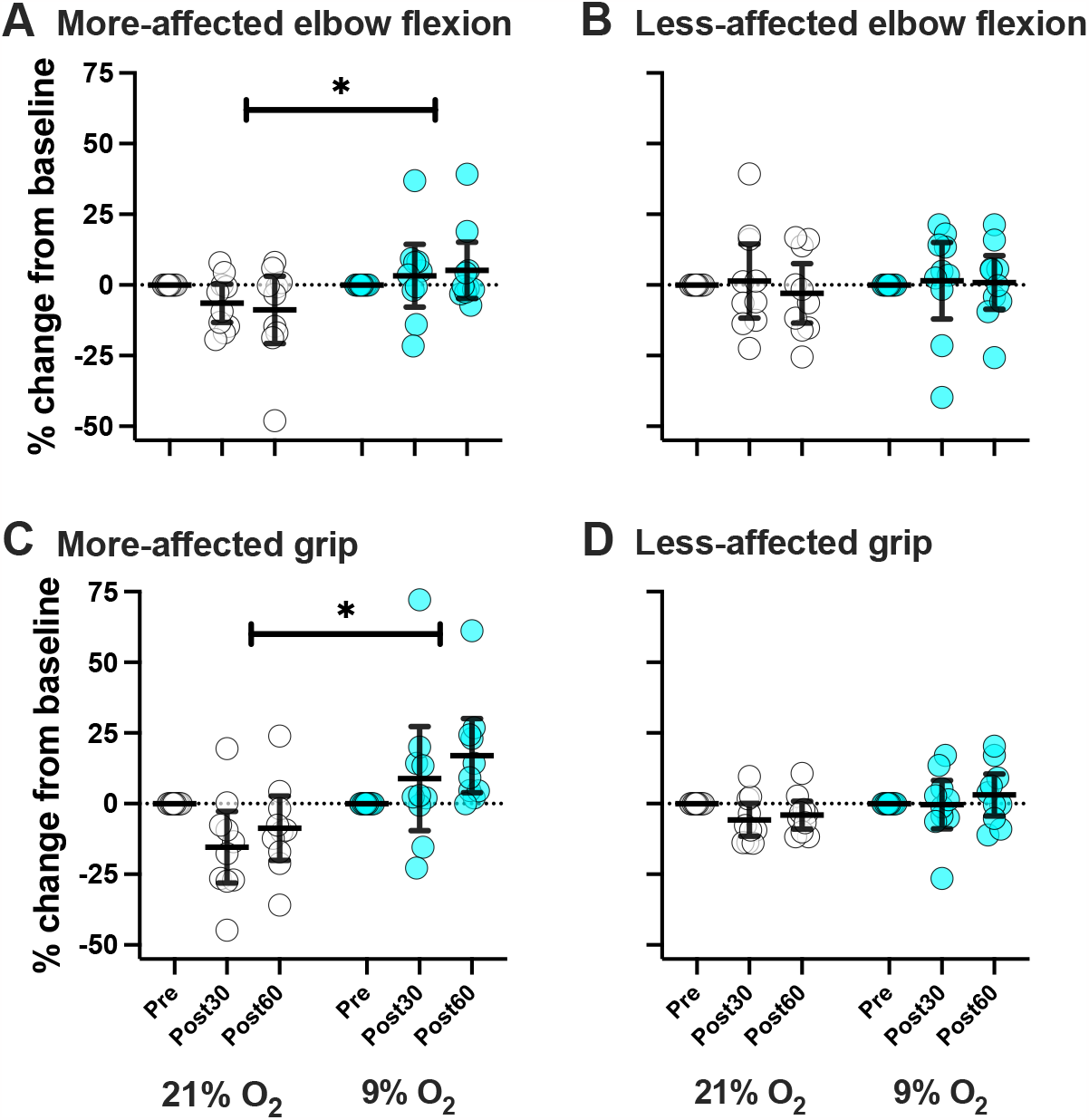
Percent change in strength. A) more-affected elbow flexion, B) less-affected elbow flexion, C) more-affected grip, and D) less-affected grip. The 21% O2 condition is white, the 9% O2 condition is cyan, error bars are 95% confidence limits, and * indicates p<0.05 for the effect of condition.

## DISCUSSION

The primary purpose of this preliminary study was to investigate the safety of AIH in chronic hemispheric stroke survivors. Our secondary objective was to determine if AIH augments volitional strength after stroke. In support of our first objective, we showed that AIH was tolerated without negative consequences. The observed ∼5 and ∼13% increase in volitional strength of the hand and elbow musculature supports the assertion proposed in our second objective, that AIH can improve volitional strength after stroke. The mechanisms underlying the increases in strength remain speculative but might include synaptic plasticity^3^, and/or improved neural drive to the motor pool^4^, which improves motor unit output^5^.

Several limitations should be acknowledged: The sample size was small, and although we did not observe any adverse events, medical monitoring will be important in future trials. Additionally, we only administered a single bout of AIH to these individuals, and the cumulative effects of hypoxia must be monitored. Repetitive AIH may enhance neuroplasticity, and thus may provide greater therapeutic benefit than a single session^2^.

## Conclusions

Our findings suggest that AIH is potentially safe and effective for improving limb function in people with stroke. Although AIH is a promising intervention, further investigation \is required to confirm safety and to maximize therapeutic outcomes.

## Data Availability

All data produced in the present study are available upon reasonable request to the authors

## Acknowledgements

Thanks to Leah O’Shea for assisting data collection and analysis, Deena Hasselballa for help with neurological assessments, and the nurses for monitoring vitals.

